# Clinical utility of Corona Virus Disease-19 serum IgG, IgM, and neutralizing antibodies and inflammatory markers

**DOI:** 10.1101/2021.01.19.21249604

**Authors:** Ernst J. Schaefer, Florence Comite, Latha Dulipsingh, Maxine Lang, Jessica Jimison, Martin M. Grajower, Nathan E. Lebowitz, Andrew S. Geller, Margaret R. Diffenderfer, Lihong He, Gary Breton, Michael L. Dansinger, Ben Saida, Chong Yuan

## Abstract

Most deaths from severe acute respiratory syndrome coronavirus-2 (SARS-CoV-2) infection occur in older subjects. We assessed age effects and clinical utility of serum SARS-CoV-2 immunoglobulin G (IgG), immunoglobulin M (IgM), and neutralizing antibodies and serum inflammatory markers. Serum IgG, IgM, and neutralizing antibody levels were measured using chemiluminescence assays from Diazyme (Poway, CA), while serum interleukin-6 (IL-6), C reactive protein (CRP), and ferritin were measured with immunoassays obtained from Roche (Indianapolis, IN). In 79,005 subjects, IgG and IgM levels were positive (≥1.0 arbitrary units [AU]/mL) in 5.29% and 3.25% of subjects, respectively. In antibody positive subjects, median IgG levels were 3.93 AU/mL if <45 years of age, 10.18 AU/mL if 45-64 years of age, and 10.85 AU/mL if ≥65 years of age (p<0.0001). In SARS-CoV-2 RNA positive cases, family members and exposed subjects (n=1,111), antibody testing was found to be valuable for case finding, and persistent IgM levels were associated with chronic symptoms. In non-hospitalized and hospitalized subjects assessed for SARS-CoV-2 RNA (n=278), median IgG levels in AU/mL were 0.05 in negative subjects (n=100), 14.83 in positive outpatients (n=129), and 30.61 in positive hospitalized patients (n=49, p<0.0001). Neutralizing antibody levels correlated significantly with IgG (r=0.875; p<0.0001). Two or more of the criteria of IL-6 ≥10 pg/mL, CRP ≥10 mg/L, and/or IgM ≥1.0 AU/mL occurred in 97.7% of inpatients versus 1.8% of outpatients (>50-fold relative risk, C statistic 0.986, p<0.0001). Our data indicate that: 1) IgG levels are significantly higher in positive older subjects, possibly to compensate for decreased cellular immunity with aging; 2) IgG levels are important for case finding in family clusters; 3) IgG levels are significantly correlated with neutralizing antibody levels; 4) persistently elevated IgM levels are associated with chronic disease; and 5) markedly elevated IL-6, hs-CRP, and/or positive IgM accurately identify SARS-CoV-2 RNA positive subjects requiring hospitalization.

## Introduction

Coronavirus disease-2019 (COVID-19) is due to severe acute respiratory syndrome coronavirus-2 (SARS-CoV-2) infection, which has caused a world-wide pandemic. The diagnosis is made by SARS-CoV-2 RNA detection in naso-pharyngeal (NP) swabs, oro-pharyngeal (OP) swabs, nasal swabs, or saliva usually within five days of exposure [1-6]. Up to 50% of SARS-CoV-2 positive patients can remain asymptomatic; however, such individuals can spread infections [7,8]. The average onset of symptoms after infection is about 5 days (range 2-14 days). Antibody testing has been reported to be useful for documenting exposure and potential immunity, as well as for case finding in family clusters and exposed individuals [9-15]. Moreover, treatment of hospitalized COVID-19 patients with convalescent plasma rich in antibodies may be useful in treating the disease [16-20].

In RNA positive subjects, IgM antibody levels are detectable within a median time of 5 days (range 3-7 days) of symptom onset and generally disappear over time, while IgG and neutralizing antibodies are detectable within a median time of 14 days (range 10-18 days) of symptom onset and generally persist for many months [9-15, 21,22]. Similar results for SARS-CoV-2 antibodies have been obtained with chemiluminescence and enzyme linked immunoassays [9-15]. Levels of IgG antibodies have been shown to correlate with levels of neutralizing antibodies in serum with some assays, but not with others [21,22]. Fingerstick antibody testing with some lateral flow devices may be unreliable [22,23]. It has been reported by the Centers for Disease Control in the United States that about 80% of the total deaths attributed to SARS-CoV-2 occur in subjects ≥65 years of age, while this group only accounts for about 10% of the total cases [24]. Our goals in the current investigation were to assess 1) the effects of age on serum SARS-CoV-2 IgG and IgM antibody levels, 2) the clinical utility of such assays in case finding and symptom prediction, 3) the relationships of IgG and IgM antibody levels with neutralizing antibody levels, and 4) the relationships of antibody levels and inflammatory markers in SARS-CoV-2 positive patients requiring hospitalization, as compared to those in positive patients not requiring hospitalization, in order to develop a risk prediction model.

## Methods

### Human subjects

A total of 79,005 subjects (median age 49.0 [IQR 35.0-69.0] years; 58.9% female, 18.2% ≥65 years of age) had serum samples submitted to our laboratory for the measurement of serum IgG levels. A subset of 62,048 subjects had samples submitted to our laboratory for the measurement of IgM levels. These subjects were assessed by healthcare providers in offices, clinics, hospitals, and at one meat packing plant (n=352). Samples were submitted to our laboratory between April 6^th^ and September 1^st^, 2020. We report data from 39 states with more than 100 results. We also assessed data from samples collected by a healthcare provider from employees at a local meat packing plant in Massachusetts (n=217). In addition, we evaluated data on samples and information about clinical status submitted by healthcare providers for 534 outpatients and selected inpatients (median age 46 years, 51.2% female) from the Boston, Bronx, Manhattan, and northern New Jersey areas. For this latter research, subject data were extracted from medical records without name or identification number and were analyzed as anonymized data.

This type of research is exempted from requirement for human institutional review board (IRB) approval as per exemption 4, as listed at https://grants.nih.gov/policy/humansubjects.htm and at the open education resource (OER) website for research involving human subjects. This exemption “involves the collection or study of data or specimens if publicly available or recorded such that subjects cannot be identified”. We had this designation and our research reviewed by the Advarra Institutional Review Board (Columbia, MD). They determined that “had the request for exempt determination been submitted prior to initiation of research activities, the research would have met the criteria for exemption from institutional review board review under 45 CFR 46.104(d)” and, therefore, ruled that this research did not require IRB approval. The anonymized data and material used for this analysis can be obtained by contacting the corresponding author (EJS) at.

We also measured serum SARS-CoV-2 IgG, IgM, and neutralizing antibodies and serum interleukin-6 (IL-6), high sensitivity C reactive protein (hs-CRP), and ferritin in 100 SARS-CoV-2 RNA negative control subjects, 129 SARS-CoV-2 RNA positive subjects not requiring hospitalization, and 49 SARS-CoV-2 RNA positive subjects requiring hospitalization (median age 48.9 years; 54.5% female; 85% Caucasian, 10 % Hispanic, and 7% African American). These subjects were enrolled in an IRB-approved protocol at Trinity Health of New England (Hartford, CT, USA); all subjects provided an informed written consent.

### SARS-CoV-2 viral detection

Detection of SARS-CoV-2 RNA in NP, OP, or nasal swabs was performed using reverse transcriptase polymerase chain reaction methods with Thermo-Fisher TaqPath COVID-19 Combo kits (Waltham, MA). This assay targets a region in the *N* gene, a region in the spike glycoprotein or *S* gene, and a region in the *ORF1* gene for SARS-CoV-2 RNA detection in swab samples. Positive values are those detected at a cycle threshold values of ≤37 cycles. Our modified version of this assay which has received emergency use authorization (EUA) from the Food and Drug Administration (FDA) was performed as previously described [4]. Our assay was found to have 100% concordance in 100 positive and 100 negative samples when compared with another RNA assay from Viracor (Lee’s Summit, MO) as previously described [5].

### SARS-CoV-2 IgG and IgM chemiluminescence assays

The assays used were the SARS-CoV-2 IgM and SARS-CoV-2 IgG chemiluminescence assays obtained from Diazyme Laboratories (Poway, CA) as previously described [10,14,15]. The assays use 2 recombinant antigens (full-length SARS-CoV-2 nucleocapsid protein and partial-length glycoprotein spike protein). The prediluted sample, buffer and magnetic microbeads coated with SARS-CoV-2 recombinant antigens are thoroughly mixed and incubated, forming immune-complexes. The precipitate is separated in a magnetic field and washed before *N*-(4-Aminobutyl)-*N*-ethyl-isoluminol labeled anti-human IgM or IgG antibodies are added and incubated to form additional complexes. After a second precipitation in a magnetic field and subsequent wash cycles, the Starter 1+2 is added to initiate a chemiluminescent reaction. The light signal is measured by a photomultiplier as relative light units (RLUs), which are proportional to the concentration of SARS-CoV-2 IgM or IgG present in the sample and are converted to arbitrary units or AU/mL.

The SARS-CoV-2 IgG antibody test did not detect SARS-CoV-2 IgM antibodies, and the SARS-CoV-2 IgM antibody test did not detect SARS-CoV-2 IgG antibodies. For cross reactivity experiments, a total of 143 clinical samples were tested with both antibody assays. These samples were confirmed positive for antibodies for various viruses and bacteria: influenza virus type A, influenza virus type B, parainfluenza virus, respiratory syncytial virus, adenovirus, EBV NA IgG, EBV VCA IgM/IgG, Measles virus, CMV IgM/IgG, Varicella zoster virus, Mycoplasma Pneumoniae IgM/IgG, Chlamydia pneumoniae IgM/IgG, Candida albicans, ANA, HCoV-HKU1, HCoV-OC43, HCoV-NL63 and HCoV-229E. All 143 samples, however, were negative for SARS-CoV-2 IgG/IgM with DZ-Lite SARS-CoV-2 IgG/IgM CLIA kits. In addition, these assays were found to have no cross reactivity with antibodies for non-SARS-CoV-2 coronavirus strains HKU1, NL63, OC43, or 229E.

Multiple serum samples with IgM concentrations ranging from 0.86-10.27 AU/mL and IgG concentrations ranging from 8.04 – 67.92 AU/mL had 0.1 mg/mL of the S protein and 0.1 mg/mL of the N protein added. After 10-minute incubations and remeasurements, mean IgM levels were reduced by 94.55% and mean IgG levels by 99.46%. These data confirmed that the antibodies measured in these assays are directed against the S and N proteins of SARS-CoV-2.

The specificity of the IgG assay for identifying 852 SARS-CoV-2 RNA negative outpatients was 97.40% when using IgG only; when used in combination with the IgM, the specificity was 96.00%. In 200 SARS-CoV-2 negative hospitalized patients, the specificity for diagnosing negative patients was 97.5% for the IgG assay alone and 96.5% for both IgM and IgG. In SARS-CoV-2 RNA positive patients (n=55), the sensitivity for detecting positive subjects for the IgG assay was 98.40% for those with symptoms ≥15 days; together with IgM it was 98.20%.

Positive values for both chemiluminescence assays are ≥1.0 AU/mL, with linear and reproducible reportable ranges of 1.0–10.0 AU/mL for IgM and of 0.20–100.00 AU/mL for IgG. Linearity studies documented r^2^ values of 0.991 for both IgM and for IgG for actual values versus target values, with within- and between-run coefficients of variation based on 20 analyses at 4 concentration levels of 4.00% and 2.51% for IgM positive (≥1.0 AU/mL) control samples and 2.50% and 2.10% for IgG positive (≥1.0 AU/mL) control samples, respectively. Both these assays have received emergency use authorization (EUA) from the Food and Drug Administration (FDA).

### Neutralizing antibody chemiluminescence assay

The SARS-CoV-2 neutralizing antibody assay utilized was obtained from Diazyme Laboratories (Poway, CA). This assay is a competitive chemiluminescence immunoassay based on the specific interaction between the SARS-CoV-2 spike protein receptor binding domain (RBD) and the human angiotensin-converting enzyme 2 receptor (hACE2) on the surface of host cells. The assay has been previously described [15]. In the absence of SARS-CoV-2 neutralizing antibodies, hACE2 and RBD form complexes that generate a high chemiluminescent signal (measured in relative light units, RLU). In the presence of SARS-CoV-2 neutralizing antibodies originating from human serum or plasma, the interaction between hACE2 and RBD is compromised; and the chemiluminescent signal is reduced in a dose-dependent manner. The assay has been validated with a cell-based assay as previously described [26]. The assay was documented to have no interfering substances and to be specific for SARS-CoV-2.

The assay showed excellent correlation with the cell-based SARS-CoV-2 Reporter Neutralizing Antibody Assay. Serum samples (n=33) with neutralizing antibody values ≥2.60 AU/mL all showed >98.0% inhibition of viral infection in cell-based assay validation studies. In our laboratory this assay was found to have within- and between-run coefficients of variation of <4.0%, with a positive value being ≥1.0 AU/mL and a linear range up to 30 AU/mL. This assay has been submitted to FDA for EUA.

### Inflammatory marker assays

Serum hs-CRP and ferritin were measured using FDA-approved assays from Roche Diagnostics (Indianapolis, IN) on a Roche c701 automated COBAS analyzer. The IL-6 immunoassay used was also obtained from Roche Diagnostics and was run on a Roche c801 automated COBAS analyzer. This assay has received FDA EUA for use in hospitalized COVID-19 patients (n=49) who are at >4-fold increased risk of needing a ventilator if their serum IL-6 values are >35 pg/mL versus patients with values ≤35 pg/mL. This information was provided in the Roche assay package insert. All assays had coefficients of variation of 4.0%.

### Statistical analysis

All statistical analyses were performed using R software, version 3.6 (R Foundation, Vienna, Austria). Categorical variables were expressed as frequencies and percentages, while continuous variables were expressed as median values with interquartile ranges (IQR, 25^th^–75^th^ percentile values). The statistical significance of differences between groups were assessed using non-parametric Kruskal-Wallis analysis. Spearman correlation analyses were performed to assess interrelations of biochemical variables. Univariate and stepwise multivariate regression analyses were carried out to assess for the statistical significance of associations.

## Results

### Antibody testing in a reference laboratory population

Table 1 shows the results of serum antibody testing at Boston Heart Diagnostics between April 6^th^ and September 1^st^, 2020 by state in which more than 100 results were reported. The highest IgG and IgM positive rates were seen for Nebraska, but these were all meat packing plant employees. New York state had a fairly low positive rate because most subjects were sampled as part of health screening. In contrast, high IgG and IgM positive rates were observed in Pennsylvania from a program that screened newly symptomatic patients.

**TABLE 1.** SARS-CoV-2 antibody testing by states with >100 tests

| State | Antibody Tests Done,<br>N | SARS-CoV-2 IgG,<br>% Positive | SARS-CoV-2 IgM,<br>% Positive |
| --- | --- | --- | --- |
| Alabama | 141 | 4.96 | 0.71 |
| Arkansas | 286 | 5.94 | 2.53 |
| California | 2,538 | 4.29 | 2.69 |
| Colorado | 187 | 6.42 | 3.40 |
| Connecticut | 1,174 | 8.43 | 2.99 |
| Florida | 1,271 | 6.06 | 3.15 |
| Georgia | 918 | 5.56 | 2.41 |
| Idaho | 184 | 2.72 | 1.64 |
| Indiana | 640 | 10.16 | 3.83 |
| Iowa | 116 | 2.72 | 1.72 |
| Massachusetts | 723 | 7.88 | 2.77 |
| Michigan | 900 | 7.67 | 3.89 |
| Missouri | 6,339 | 14.59 | 11.25 |
| Nebraska* | 352 | 19.03 | 15.34 |
| Nevada | 183 | 3.83 | 0.56 |
| New Jersey | 284 | 13.73 | 7.42 |
| New York† | 57,985 | 4.70 | 2.81 |
| North Carolina | 469 | 2.35 | 1.94 |
| Ohio | 139 | 1.44 | 2.17 |
| Oklahoma | 228 | 11.84 | 2.28 |
| Oregon | 1,346 | 2.60 | 3.60 |
| Pennsylvania‡ | 169 | 11.83 | 13.61 |
| Texas | 1,343 | 4.02 | 3.14 |
| Virginia | 155 | 4.52 | 7.23 |
| Washington | 1,033 | 4.16 | 2.32 |

\*Meat packing plant.
†Mainly health screening.
‡Newly symptomatic screening program.

Table 2 shows the results of testing 79,005 subjects for IgG and a subset of 62,048 subjects that also had serum IgM levels measured. IgG and IgM levels were positive (≥1.0 AU/mL) in 5.29% and 3.25% of these subjects, respectively. In positive subjects, median IgG levels were 3.93 AU/mL if <45 years, 10.18 AU/mL if 45-64 years, and 10.85 AU/mL if ≥65 years (p<0.0001). Therefore, subjects in the ≥65-year age group and the 45-64-year group had more than two-fold higher median IgG levels than subjects <45 years of age. These findings were true for both females and males. In addition, females in the ≥65-year age group were significantly (p<0.0001) more likely to have positive IgM values than females in the <45-year age group. In addition to age effects, we also noted that men in the 45-64-year age group had significantly (p<0.001) higher IgG levels than their female counterparts.

**TABLE 2.**
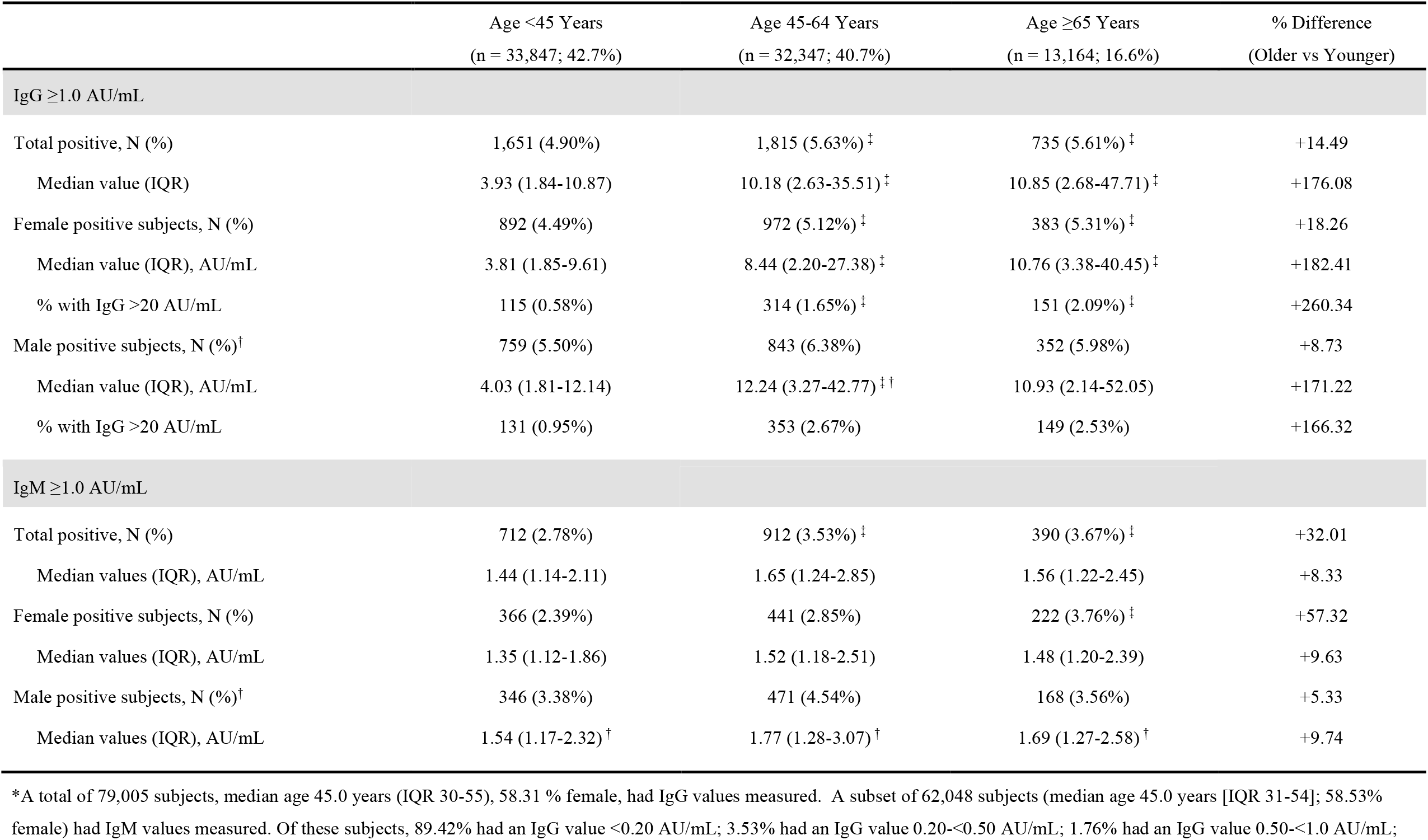
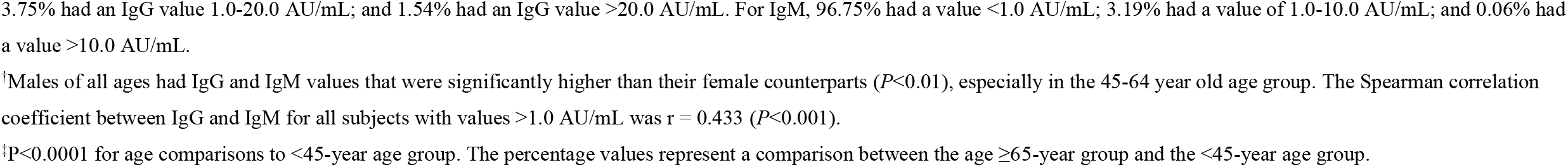
SARS-CoV-2 antibody levels by age and gender∗

|  | Age <45 Years<br>(n = 33,847; 42.7%) | Age 45-64 Years<br>(n = 32,347; 40.7%) | Age ≥65 Years<br>(n = 13,164; 16.6%) | % Difference<br>(Older vs Younger) |
| --- | --- | --- | --- | --- |
| **IgG ≥1.0 AU/mL** |  |  |  |  |
| Total positive, N (%) | 1,651 (4.90%) | 1,815 (5.63%)<sup>‡</sup> | 735 (5.61%)<sup>‡</sup> | +14.49 |
| Median value (IQR) | 3.93 (1.84-10.87) | 10.18 (2.63-35.51)<sup>‡</sup> | 10.85 (2.68-47.71)<sup>‡</sup> | +176.08 |
| Female positive subjects, N (%) | 892 (4.49%) | 972 (5.12%)<sup>‡</sup> | 383 (5.31%)<sup>‡</sup> | +18.26 |
| Median value (IQR), AU/mL | 3.81 (1.85-9.61) | 8.44 (2.20-27.38)<sup>‡</sup> | 10.76 (3.38-40.45)<sup>‡</sup> | +182.41 |
| % with IgG >20 AU/mL | 115 (0.58%) | 314 (1.65%)<sup>‡</sup> | 151 (2.09%)<sup>‡</sup> | +260.34 |
| Male positive subjects, N (%)<sup>†</sup> | 759 (5.50%) | 843 (6.38%) | 352 (5.98%) | +8.73 |
| Median value (IQR), AU/mL | 4.03 (1.81-12.14) | 12.24 (3.27-42.77)<sup>‡†</sup> | 10.93 (2.14-52.05) | +171.22 |
| % with IgG >20 AU/mL | 131 (0.95%) | 353 (2.67%) | 149 (2.53%) | +166.32 |
| **IgM ≥1.0 AU/mL** |  |  |  |  |
| Total positive, N (%) | 712 (2.78%) | 912 (3.53%)<sup>‡</sup> | 390 (3.67%)<sup>‡</sup> | +32.01 |
| Median values (IQR), AU/mL | 1.44 (1.14-2.11) | 1.65 (1.24-2.85) | 1.56 (1.22-2.45) | +8.33 |
| Female positive subjects, N (%) | 366 (2.39%) | 441 (2.85%) | 222 (3.76%)<sup>‡</sup> | +57.32 |
| Median values (IQR), AU/mL | 1.35 (1.12-1.86) | 1.52 (1.18-2.51) | 1.48 (1.20-2.39) | +9.63 |
| Male positive subjects, N (%)<sup>†</sup> | 346 (3.38%) | 471 (4.54%) | 168 (3.56%) | +5.33 |
| Median values (IQR), AU/mL | 1.54 (1.17-2.32)<sup>†</sup> | 1.77 (1.28-3.07)<sup>†</sup> | 1.69 (1.27-2.58)<sup>†</sup> | +9.74 |

\*A total of 79,005 subjects, median age 45.0 years (IQR 30-55), 58.31 % female, had IgG values measured. A subset of 62,048 subjects (median age 45.0 years [IQR 31-54]; 58.53% female) had IgM values measured. Of these subjects, 89.42% had an IgG value <0.20 AU/mL; 3.53% had an IgG value 0.20-<0.50 AU/mL; 1.76% had an IgG value 0.50-<1.0 AU/mL;
3.75% had an IgG value 1.0-20.0 AU/mL; and 1.54% had an IgG value >20.0 AU/mL. For IgM, 96.75% had a value <1.0 AU/mL; 3.19% had a value of 1.0-10.0 AU/mL; and 0.06% had a value >10.0 AU/mL.
<sup>†</sup>Males of all ages had IgG and IgM values that were significantly higher than their female counterparts ( $P<0.01$ ), especially in the 45-64 year old age group. The Spearman correlation coefficient between IgG and IgM for all subjects with values >1.0 AU/mL was $r = 0.433$ ( $P<0.001$ ).
<sup>‡</sup> $P<0.0001$ for age comparisons to <45-year age group. The percentage values represent a comparison between the age $\geq 65$ -year group and the <45-year age group.

### Studies in meat processing plants

Of 352 subjects from a meat packing plant in Nebraska (Table 1), 19.0% had positive IgG values; and 15.3% had positive IgM values. In a separate analysis by one of our healthcare providers of 217 employees at a local meat processing plant in Massachusetts tested with NP swabs, 24.0% were RNA positive. When 41 of these 52 positive subjects were retested 2 weeks later, 73.2% still had positive NP swabs, 70.7% had positive IgG values, 9.8% had positive IgM values, and 63.4% had been symptomatic. Median IgG and IgM in all 41 subjects tested were 20.53 AU/mL and 0.54 AU/mL, respectively. As shown in Figure 1A, there was a very large variability in IgG response (range <0.20-117.7 AU/mL). In addition, there were 25 subjects that had prior RNA negative swab testing, but requested antibody testing because of having significant symptoms and known exposure to subjects that had tested positive with RNA testing. Of these, 64.0% had positive IgG levels and 28.0% had positive IgM values, with all subjects in the latter group having persistent symptoms. Median IgG and IgM values in these positive subjects were 24.73 AU/mL and 1.31 AU/mL, respectively. These data clearly document the benefits of antibody testing for case finding in previously exposed subjects even with negative RNA tests.

**FIGURE 1.**
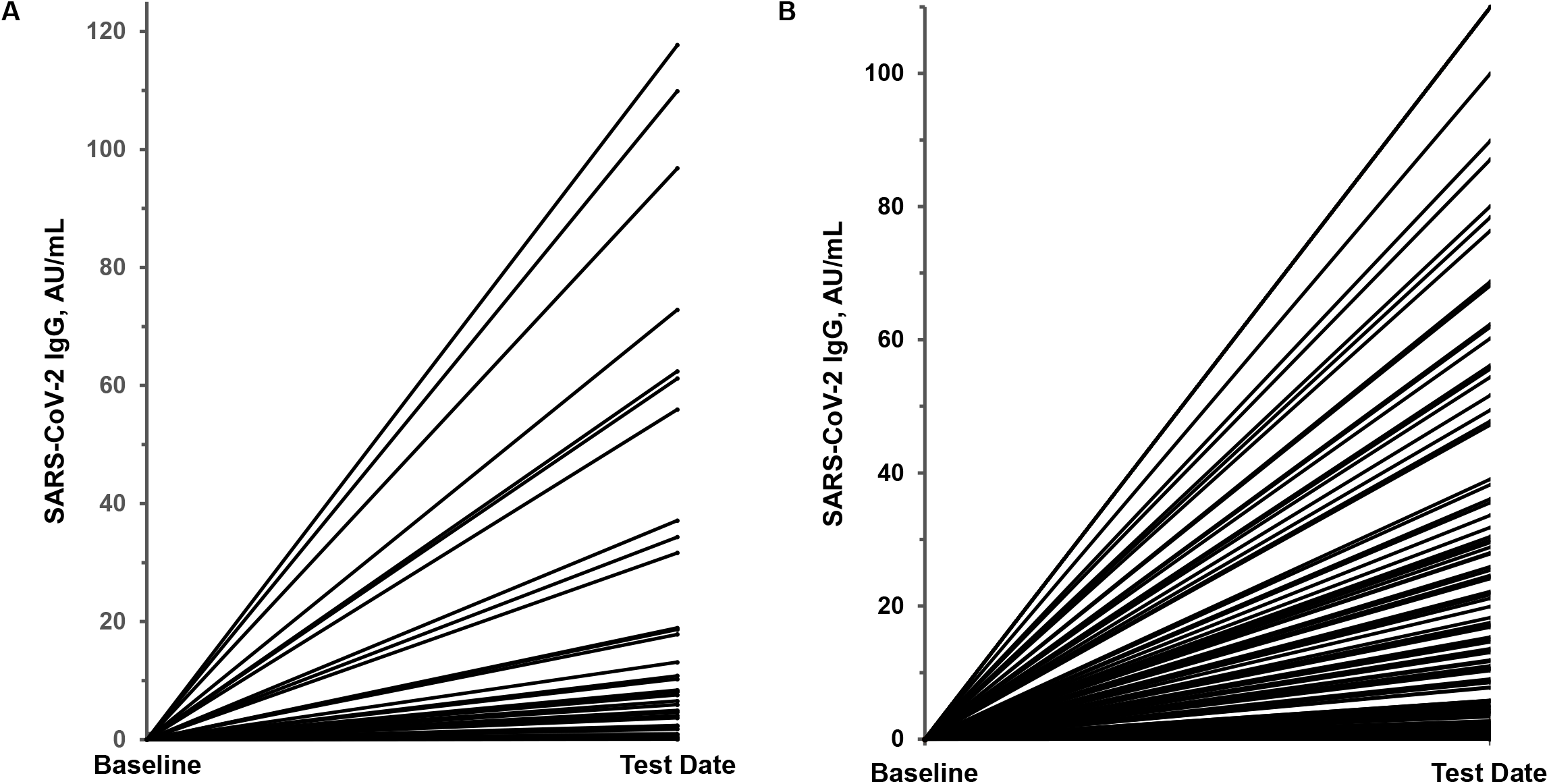
Variability in SARS-CoV-2 IgG antibody response. Panel A shows the response in 41 meat packing plant employees who were SARS-CoV-2 RNA positive 2 weeks prior to testing; panel B, the response in 129 potential convalescent plasma donors 4-6 weeks after a SARS-CoV-2 RNA positive test. The baseline value was assumed to be IgG 0.05 AU/mL.

### Studies in healthcare provider’s office

Of 388 outpatients that had antibody testing in a healthcare provider’s office (MMG) in the Riverdale area of the Bronx, NY, 17.5% had positive IgG values with or without positive IgM values, while another 4.9% had borderline IgG values between 0.50-1.0 AU/mL. Of these latter subjects, 60.0% had been or were symptomatic. Of 10 subjects in the borderline category, 3 were previously RNA positive on swabs, and 6 had a history of definite exposure. This healthcare provider felt that IgG values between 0.50-1.0 AU/mL should be classified as borderline. His data justified this conclusion.

### Antibody levels in individual family clusters and cases

Of 154 outpatients in Manhattan and New Jersey that had NP swabs and antibodies assessed, 85.8% were negative for any evidence of SARS-CoV-2. The remaining 14.2% (n=22) were positive and of these subjects, 7 were carefully followed over time along with their family members, as well as 9 individual cases (total of 47 subjects). Many had the following symptoms: fever, chills, body aches, inability to sleep, fatigue, dry cough, loss of smell and taste, shortness of breath, and diarrhea. Three cases (all aged >80 years) had to be hospitalized, and two required being placed on ventilators, with one of these latter patients dying. The data that we tabulated clearly indicated that 1) antibody testing was valuable for finding additional cases in family studies (observed in all families); 2) patients can have positive RNA results for up to 6 weeks (observed in 5 cases); and 3) patients with persistent symptoms often have persistently elevated IgM levels (observed in 11 cases).

### Antibody and inflammation biomarker levels in RNA positive outpatients and inpatients

Data on serum IgG, IgM, and neutralizing antibody levels, as well as the inflammatory markers IL-6, hsCRP, and ferritin, in 100 SARS-CoV-2 RNA negative control subjects, 129 SARS-CoV-2 RNA positive outpatients, and 49 SARS-CoV-2 RNA positive inpatients are shown in Table 3. All control subjects had negative antibody levels (<1.0 AU/mL). Median IgG levels were about 300-fold and 600-fold higher in outpatients and inpatients as compared to controls (both p<0.0001). The wide variation in IgG response in outpatients is shown in Figure 1B. IgG values ranged from 1.03-200.0 AU/mL in outpatients and from 0.05-169.5 AU/mL in inpatients, similar to what we observed in positive meat packing plant employees (Figure 1A). Median IgM levels were about 1.8-fold and 5-fold higher in outpatients and inpatients as compared to controls (both p<0.0001). IgM values ranged from 1.09-13.58 AU/mL in outpatients and from 0.46-18.82 AU/mL in inpatients. Median neutralizing antibody levels were about 12-fold and 24-fold higher in outpatients and inpatients as compared to controls (both p<0.0001). Neutralizing antibody values ranged from 1.09-13.58 AU/mL in outpatients and from 0.35-18.82 AU/mL in inpatients. Median IgG, IgM, and neutralizing antibody levels were all significantly higher in positive patients than controls.

**TABLE 3.**
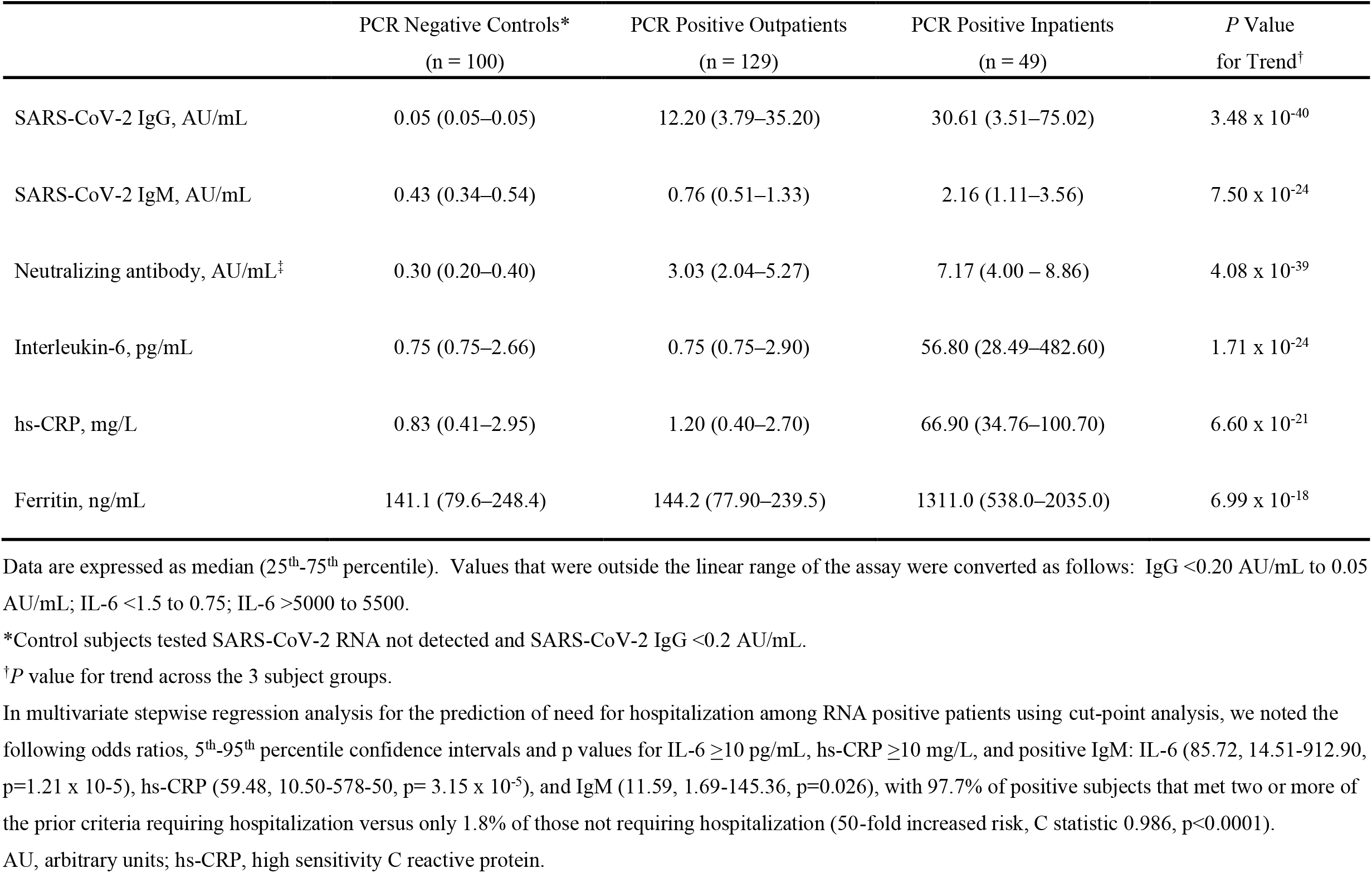
Antibody and inflammatory biomarker response in SARS-CoV-2 PCR positive outpatients and PCR positive inpatients vs PCR negative control subjects

Median IL-6 levels were the same in controls and outpatients but were about 75-fold higher in inpatients as compared to other groups (p<0.0001). IL-6 values ranged from 0.75-39.42 pg/mL in controls, from 0.75-42.30 pg/mL in outpatients, and from 0.75-5500 pg/mL in inpatients. Median hs-CRP levels were very similar in control subjects and outpatients but were about 80-fold higher in inpatients as compared to other groups (p<0.0001). Hs-CRP values ranged from 0.06-63.10 mg/L in controls, from 0.15-73.10 mg/L in outpatients, and from 1.22-375.60 mg/L in inpatients. Similarly, median ferritin levels were similar in controls and outpatients, but were about 9-fold higher in inpatients as compared to other groups (p<0.0001). Ferritin values ranged from 9.0-1413.0 ng/mL in controls, from 14.0-786.4 ng/mL in outpatients, and from 130.0-2653.0 ng/mL in inpatients (p<0.0001). Levels of inflammatory markers were only significantly elevated in inpatients as compared to controls and outpatients. Using widely measured hs-CRP values and a cut-point of ≥10 mg/L, 6.0% of negative control subjects, 5.6% of positive outpatients, and 93.2% of positive inpatients had values above this threshold. Therefore, RNA positive subjects with hs-CRP values ≥10 mg/L had a 16.6-fold higher risk (p<0.0001) of being hospitalized than RNA positive subjects with hs-CRP values <10 mg/L.

Correlation matrix data for levels of antibodies and inflammation markers are shown in Table 4. Neutralizing antibody levels correlated most significantly (p<0.0001) with IgG (r=0.875) and IgM (r=0.654), while IL-6 values correlated most strongly with hs-CRP values (r=0.740, p<0.0001). We also sought to develop a multi-parameter algorithm to distinguish RNA positive subjects who required hospitalization from RNA positive subjects not requiring hospitalization. In multivariate stepwise regression analysis for the prediction of need for hospitalization among RNA positive patients using cut-point analysis, we noted the following odds ratios (5^th^-95^th^ percentile confidence intervals; p values) for IL-6 ≥10 pg/mL, hs-CRP ≥10 mg/L, and/or IgM ≥1.0 AU/mL: IL-6 85.72 (CI, 14.51-912.90; p=1.21 x 10^−5^), hs-CRP 59.48 (CI 10.50-578-50; p= 3.15 x 10^−5^), and IgM 11.59 (CI 1.69-145.36; p=0.026), with 97.7% of positive subjects that met two or more of the prior criteria requiring hospitalization versus only 1.8% of those not requiring hospitalization (50-fold increased risk, C statistic 0.986, p<0.0001). Neither ferritin, IgG, or neutralizing antibody values added significant risk prediction to this model after the first three variables had been entered.

**TABLE 4.**
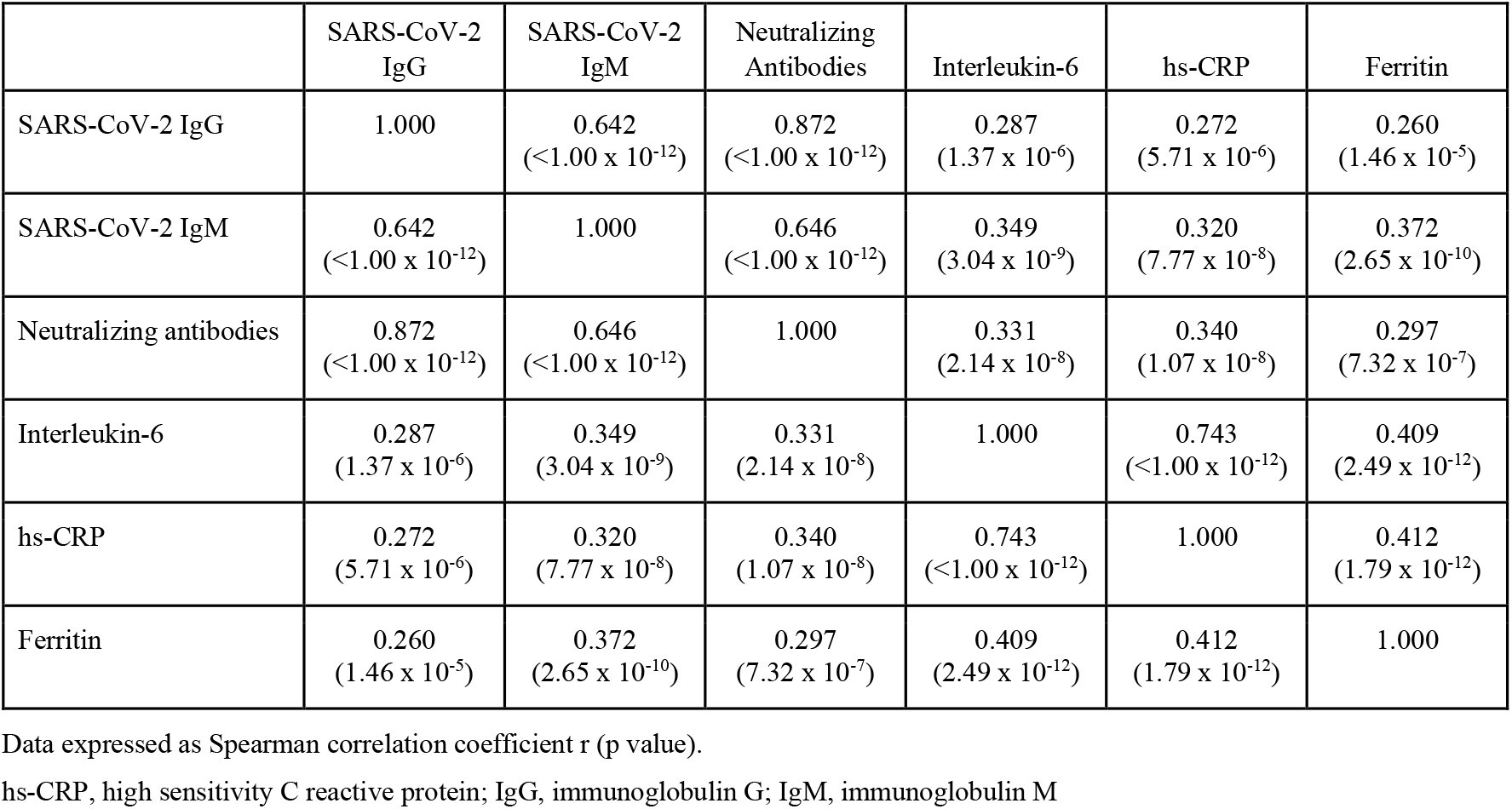
Spearman correlation coefficient matrix analysis of antibody and inflammatory marker response in all subjects (n = 278)

## Discussion

One of our goals in the current investigation was to assess the effects of age on serum SARS-CoV-2 IgG and IgM antibody levels. In a large number of outpatients with potential SARS-CoV-2 exposure, about 5% had positive IgG values and about 3% had positive IgM values. We noted significant age effects, with positive subjects ≥65 years and those between 45-64 years of age having median IgG levels that were more than two-fold higher than their younger counterparts. We also noted a modest sex effect with men in the 45-64-year age group having significantly higher IgG levels than their female counterparts. It has been reported by the Centers for Disease Control and Prevention (CDC) that serum SARS-CoV-2 antibody levels were positive in 1.0-6.5% of 16,025 subjects in various parts of the United States, suggesting that infection rates were 6-24 times higher than reported at that time [26]. These percentages are similar to our data. Based on CDC data, over 95% of deaths from COVID occur in the >45-year age group, even though about 70% of the cases occur in those <45 years of age. The ≥65 years of age category accounts for ∼10% of all SARS-CoV-2 cases and ∼80% of SARS-CoV-2 mortality [24]. Possibly older subjects with positive antibody levels mount a greater IgG response in order to compensate for the decreased overall cellular immunity found in the elderly as compared to the young [28,29].

A second goal of our studies was to assess the clinical utility of antibody assays in case finding. We documented that antibody testing was valuable to identify cases and to ascertain potential exposure and level of immunity. The latter findings are relevant in the identification of potential convalescent plasma donors to assure sufficient IgG antibody levels. We have noted a high degree of variability in IgG antibody response in RNA positive patients. Laboratories that only report a positive or negative value do not detect this large variability. Moreover, only about 50% of RNA positive outpatients had IgG levels >6.5 AU/mL, sufficient to provide estimated antibody titers of >1:320 as per FDA guidance, and only about one-third had plasma IgG levels >20 AU/mL, sufficient to provide estimated antibody titers >1:1000 [16-20].

In our individual and cluster studies, we have noted that antibody testing allows for the identification of exposed individuals, especially in those that were negative based on NP swab testing, usually ≥4 weeks following infection. Most of these family cluster and individual cases studies were carried by one of the co-authors (FC). She justifiably emphasized the value of both RNA and antibody testing in her practice. Her data clearly documented the benefits of semi-quantitative IgG and IgM testing for case finding in family clusters and exposed subjects who were RNA negative. Her data also indicated that RNA swabs can remain positive for up to 6 weeks, even though such patients may no longer be able to infect other people [30,31]. In her cluster and case data, we also clearly observed that long-term elevated IgM levels were often associated with persistent illness and symptoms. At the present time, very few healthcare providers are measuring COVID-19 antibody levels; instead, there has been a frenzy of nasal swab RNA testing [25]. Unfortunately, such testing in the United States has often been accompanied by a lack of public health measures as well as contact tracing to combat the spread of COVID-19. In our view antibody testing provides an excellent measure of prior exposure and potential immunity that has been greatly under-utilized in the United States [32].

A third goal of our studies was to investigate the relationships of neutralizing antibody levels with IgG and IgM antibody levels. We noted that both IgG and IgM were significantly correlated with neutralizing antibodies, but this relationship was strongest with IgG, consistent with prior reports [21]. A great advantage of the serum or plasma neutralizing assay we used in our studies was its ease of use on high through-put automated instruments and its reproducibility. Moreover, the results of this assay were found to be very highly correlated with results obtained using a cell-based assay [26]. The utility of antibody testing after vaccination has not been assessed.

A fourth goal of our studies was to examine the relationships of antibody levels and inflammatory markers in SARS-CoV-2 positive patients requiring hospitalization as compared to such subjects not requiring hospitalization in order to potentially develop a risk algorithm. The highest median IgG, IgM, and neutralizing antibody levels that we observed were noted in hospitalized COVID-19 patients. As we have previously noted there was a high degree of variability in IgG response (Figure 1). In our multivariate analysis, three parameters allowed for the prediction of the need for hospitalization in RNA positive patients: IL-6 ≥10.0 pg/mL, hs-CRP ≥10 mg/L, and/or IgM ≥1.0 AU/mL. IL-6 and hs-CRP have been part of so called “cytokine storm” associated with an exaggerated immune response with markedly elevated blood levels of white blood cells, CRP, IL-6, and ferritin associated with a high COVID-19 mortality [33,34]. In a meta-analysis, IL-6 levels were reported to be >12-fold elevated in COVID-19 related respiratory distress [35]. Moreover, serum levels of IL-6 >80 pg/mL and hs-CRP >97 mg/L have been reported to identify correctly 80% of hospitalized COVID-19 patients requiring a ventilator with AUC values of 0.90 and 0.97, respectively [36]. In our data set, having hs-CRP value >10 mg/L increased hospitalization risk 16-fold rate in positive subjects. However, having two or more of the criteria of a positive IL-6 ≥10 pg/mL, hs-CRP ≥10 mg/L, and an IgM ≥1.0 AU/mL increased the hospitalization risk >50-fold in COVID positive patients with an AUC value of 0.98. Therefore, using these serum markers, one can very accurately predict need for hospitalization among SARS-CoV-2 RNA positive patients.

## Conclusions

Our data are consistent with the following conclusions: 1) SARS-CoV-2 IgG antibody levels are significantly higher in positive older subjects than in younger positive subjects possibly to compensate for the decreased cellular immunity observed in older people; 2) elevated SARS-CoV-2 IgG levels measurements are useful in identifying cases in exposed subjects and family clusters, 3) elevated SARS-CoV-2 IgM levels are often associated with persistent COVID-19 symptoms and disease; 4) serum SARS-CoV-2 IgG antibody levels are significantly correlated with neutralizing antibody levels; and 5) having two or more of the following laboratory abnormalities IL-6 ≥10 pg/mL, hs-CRP ≥10.0 mg/L, and/or IgM ≥1.0 AU/mL very accurately predicts the need for hospitalization in COVID-19 positive patients.

## Data Availability

The raw data from this study are available upon request to the corresponding author.

## Acknowledgements

We thank the laboratory staff at Boston Heart Diagnostics, Framingham, MA, and Diazyme Laboratories, Poway, CA, and the clinical staff at the Comite Center for Precision Medicine and Health, New York, NY, St. Francis Hospital/Trinity Health of New England, Hartford, CT, Atkinson Family Practice, Amherst, MA, Grajower Clinical Practice, the Bronx, NY, and the Advanced Cardiology Institute, Fort Lee, NJ for their efforts and commitment to SARS-CoV-2 testing, diagnosis, and treatment.

## References

1. Zhou, X.-L. Yang, X.-G. Wang, B et al. A pneumonia outbreak associated with a new coronavirus of probable bat origin. Nature 2020;579:270–273. doi:10.1038/s41586-020-2012-7 [published online first: February 3, 2020].

2. Zhu N, Zhang D, Wang W et al. A novel coronavirus from patients with pneumonia in China, 2019. N Engl. J Med 2020;382:727–733. doi:10.1056/NEJMoa2001017 [published online first: January 24, 2020].

3. Wylie AL, Fournier J, Casanovas-Massana A et al. Saliva or nasopharyngeal swab specimens for detection of SARS-CoV-2. N Engl J Med 2020;383:1283–1286. doi:10.1056/NEJMc2016359 [published online first: August 28, 2020].

4. Kleiboeker S, Cowden S, Grantham J et al. SARS-CoV-2 viral load assessment in respiratory samples. J Clin Virol 2020;129:104439. doi:10.1016/j.jcv.2020.104439 [published onfline first: May 19, 2020].

5. Schaefer EJ, Geller AS, Diffenderfer MR et al. Variability in coronavirus disease-2019 case, death, and testing rates in the United States and worldwide. medRxiv doi:10.1101/2020.10.13.20172957v2 [posted December 31, 2020].

6. Wiersinga WJ, Rhodes A, Cheng AC, Peacock SJ, Prescott HC. Pathophysiology, transmission, diagnosis, and treatment of Coronavirus Disease 2019 (COVID-19): A review. JAMA 2020;324:782–93. doi:10.1001/jama.2020.12839.

7. Rothe C, Schunk M, Sothmann P et al. Transmission of 2019-nCoV Infection from an asymptomatic contact in Germany. N Engl J Med 2020;382:970–971. doi:10.1056/NEJMc2001468 [published online first: January 30, 2020].

8. Mizumoto K, Kagaya K, Zarebski A, Chowell G. Estimating the asymptomatic proportion of coronavirus disease 2019 (COVID-19) cases on board the Diamond Princess cruise ship, Yokohama, Japan, 2020. Euro Surveill 2020;25:2000180. doi:10.2807/1560-7917.ES.2020.25.10.2000180.

9. Guo L, Ren L, Yang S et al. Profiling early humoral response to diagnose novel coronavirus disease (COVID-19). Clin Infect Dis 2020;71:778–785. doi:10.1093/cid/ciaa310.

10. Lippi G, Salvagno GL, Pegoraro M et al. Assessment of immune response to SARS-CoV-2 with fully automated MAGLUMI 2019-nCoV IgG and IgM chemiluminescence immunoassays. Clin Chem Lab Med 2020; 58:1156-1159. doi.10.1515/cclm-2020-0473 [published online first: April 16, 2020].

11. Okba NMA, Müller MA, Li W et al. Severe acute respiratory syndrome coronavirus 2−specific antibody responses in coronavirus disease patients. Emerg Infect Dis 2020;26:1478–1488. doi:10.3201/eid2607.200841 [published online first: June 21, 2020].

12. Ren L, Zhang L, Chang D et al. The kinetics of humoral response and its relationship with the disease severity in COVID-19. Commun Biol 2020;3:780. doi:10.1038/s42003-020-01526-8.

13. To KK, Tsang OT, Leung WS et al. Temporal profiles of viral load, in posterior oropharyngeal saliva samples and serum antibody responses during infection by SARS-CoV-2: an observational cohort study. Lancet Infect Dis 2020;20:565–574. doi:10.1016/51473-3099(20)30196-1 [published online first: March 23, 2020].

14. Suhandynata RT, Hoffman MA, Kelner MJ et al. Longitudinal monitoring of SARS-CoV-2 IgM and IgG seropositivity to detect COVID-19. J Appl Lab Med 2020;5:908–920. doi:10.1092/jalm/jfaa079.

15. Suhandynata RT, Hoffman MA, Kelner MJ et al. Multi-platform comparison of SARS-CoV-2 serology assays for the detection of COVID-19. J Appl Lab Med 2020;5:1324–1336. doi:10.1093/jalm/jfaa139.

16. Shen C, Wang Z, Zhao F et al. Treatment of 5 critically ill patients with COVID-19 with convalescent plasma. JAMA 2020;323:1582–1589. doi:10.1001/jama.2020.4783.

17. Li L, Zhang W, Hu Y, et al. Effect of convalescent plasma therapy on time to clinical improvement in patients with severe and life-threatening COVID-19: a randomized clinical trial. JAMA 2020;324:460–470. doi:10.1001/jama.2020.10044.

18. Dulipsingh L, Ibrahim D, Schaefer EJ et al. SARS-CoV-2 serology and virology trends in donors and recipients of convalescent plasma. Transfus Apher Sci 2020;59:102922. doi:10.1016/j.transci.2020.102922 [published online first: August 25, 2020].

19. Ibrahim D, Dulipsingh L, Zapatka L, et al. Factors associated with good patient outcomes following convalescent plasma in COVID-19: a prospective phase II clinical trial. Infect Dis Ther 2020:9:913–926. doi:10.1007/s40121-020-00341-2 [published online first: September 20, 2020].

20. CBER. Investigational COVID-19 Convalescent Plasma - Emergency INDs. https://www.fda.gov/vaccines-blood-biologics/investigational-new-drug-ind-or-device-exemption-ideprocess-cber/investigational-covid-19-convalescent-plasma-emergency-inds.

21. Wajnberg A, Amanat F, Firpo A et al. Robust neutralizing antibodies to SARS-CoV-2 infection persist for months. Science 2020;370:1227–1230. doi:10.1126/science.abd7728 [published online first: October 28, 2020].

22. Tang MS, Case JB, Franks CE et al. Association between SARS-CoV-2 neutralizing antibodies and commercial serological assays. Clin Chem 2020;66:12:1538–1547. doi:10.1093/clinchem/hvaa211 [published online first: October 7, 2020].

23. National COVID Testing Scientific Advisory Panel. Evaluation of antibody testing for SARS-CoV-2 using ELISA and lateral flow immunoassays medRxiv doi:10.1101/2020.04.15.20066407.

24. Deeks JJ, Dinnes J, Takwoingi Y et al. Antibody tests for identification of current and past infection with SARS-CoV-2. Cochrane Database Syst Rev 2020;6:CD013652. doi:10.1002/14651858.CD013652.

25. https://www.cdc.gov/covid-data-tracker/index.html#testing

26. Muruato AE, Fontes-Garfias CR, Ren P et al. A high-throughput neutralizing antibody assay for COVID-19 diagnosis and vaccine evaluation. Nat Commun 2020;11:4059. doi:10.1038/s41467-020-17892-0.

27. Havers FP, Reed C, Lim T et al. Seroprevalence of antibodies to SARS-CoV-2 in 10 sites in the United States, March 23-May 12, 2020. JAMA published online first: July 21, 2020. doi:10.1001/jamainternmed.2020.4130.

28. Wong GCL, Strickland MC, Larbi A. Changes in T cell homeostasis and vaccine responses in old age. Interdiscip Top Gerontol Geriatr 2020;43:36–55. doi:10.1159/000504487 [published online first: April 9, 2020].

29. Aspinall R. Age-related changes in the function of T cells. Microsc Res Tech 2003;62:508–513. doi:10.1002/jemt.10412.

30. He X, Lau EHY, Wu P et al. Temporal dynamics in viral shedding and transmissibility of COVID-19. Nat Med 2020;26:672–675 doi:10.1038/s41591-020-0869-5 [published online first: April 15, 2020].

31. Wölfel R, Corman VM, Guggemos W et al. Virological assessment of hospitalized patients with COVID-19. Nature 2020;581:465–469. doi:10.1038/s41586-020-2196-x [published online first: April 1, 2020].

32. Weitz JS, Beckett SJ, Coenen AR et al. Modeling shield immunity to reduce COVID-19 epidemic spread. Nat Med 2020;26:849–854. doi:10.1038/s41591-020-0895-3 [published online first: May 7, 2020].

33. Chen G, Wu D, Guo W et al. Clinical and immunologic features in severe and moderate coronavirus disease 2019. J Clin Invest 2020;130:2620-2629. doi.org/10.1172/JCI137244.

34. Ruan Q, Yang K, Wang W, Jiang L, Song J. Clinical predictors of mortality due to COVID-19 based on an analysis of data of 150 patients from Wuhan, China. Intensive Care Med 2020;46:846–848. doi:10.1007/s00134-020-05991-x [published online first: March 3, 2020].

35. Leisman DE, Ronner L, Pinotti R et al. Cytokine elevation in severe and critical COVID-19: a rapid systematic review, meta-analysis, and comparison with other inflammatory syndromes. Lancet Respir Med 2020;8:1233–1244. doi:10.1016/S2213-2600(20)30404-5 [published online first: October 16, 2020].

36. Herold T, Jurinovic V, Arnreich C et al. Elevated levels of IL-6 and CRP predict the need for mechanical ventilation in COVID-19. J Allergy Clin Immunol 2020;146:128-136.e4. doi:10.1016/j.jaci.2020.05.008 [published online first: May 18, 2020].

